# Hydroxyurea maintains working memory function in pediatric sickle cell disease

**DOI:** 10.1101/2023.11.23.23298960

**Authors:** Jesyin Lai, Ping Zou, Josue L. Dalboni da Rocha, Andrew M. Heitzer, Tushar Patni, Yimei Li, Matthew A. Scoggins, Akshay Sharma, Winfred C. Wang, Kathleen J. Helton, Ranganatha Sitaram

## Abstract

Pediatric patients with sickle cell disease (SCD) have decreased oxygen-carrying capacity in the blood and reduced or restricted cerebral blood flow resulting in neurocognitive deficits and cerebral infarcts. The standard treatment for children with SCD is hydroxyurea; however, the treatment-related neurocognitive effects are unclear. A key area of impairment in SCD is working memory, which is implicated in other cognitive and academic skills. N-back tasks are commonly used to investigate neural correlates of working memory. We analyzed functional magnetic resonance imaging (fMRI) of patients with SCD while they performed n-back tasks by assessing the blood-oxygenation level-dependent (BOLD) signals during working memory processing. Twenty hydroxyurea-treated and 11 control pediatric patients with SCD (7–18 years old) performed 0-, 1-, and 2-back tasks at 2 time points, once before hydroxyurea treatment (baseline) and ∼1 year after treatment (follow-up). Neurocognitive measures (e.g., verbal comprehension, processing speed, full-scale intelligence quotient, etc.) were assessed at both time points. Although no significant changes in behavior performance of n-back tasks and neurocognitive measures were observed in the treated group, we observed a treatment-by-time interaction in the right cuneus and angular gyrus for the 2-> 0-back contrast. Through searchlight-pattern classifications in the treated and control groups to identify changes in brain activation between time points during the 2-back task, we found more brain areas, especially the posterior region, with changes in the pattern and magnitude of BOLD signals in the control group compared to the treated group. In the control group, increases in 2-back BOLD signals were observed in the right crus I cerebellum, right inferior parietal lobe, right inferior temporal lobe, right angular gyrus, left cuneus and left middle frontal gyrus at 1-year follow-up. Moreover, BOLD signals elevated as the working memory load increased from 0- to 1-back but did not increase further from 1- to 2-back in the right inferior temporal lobe, right angular gyrus, and right superior frontal gyrus. These observations may result from increased cognitive effort during working memory processing with no hydroxyurea treatment. In contrast, we found fewer changes in the pattern and magnitude of BOLD signals across time points in the treated group. Furthermore, BOLD signals in the left crus I cerebellum, right angular gyrus, left cuneus and right superior frontal gyrus of the treated group increased continuously with increasing working memory load from 0- to 2-back, potentially related to a broader dynamic range in response to task difficulty and cognitive effort. Collectively, these findings suggest that hydroxyurea treatment helped maintain working memory function in SCD.

## Introduction

Sickle cell disease (SCD) is a devastating hematologic disease marked by acute and chronic cerebrovascular changes. Patients with SCD often have symptoms of brain injury, including neurocognitive deficits (Brandling-Bennett et al., 2003; Pavlakis SG et al., 1989). Approximately 11% of patients who have the most severe form of SCD [i.e., hemoglobin (Hb) SS] experience overt stroke before the age of 20 years (3). Moreover, 17-22% of patients with SCD experience silent cerebral infarction (SCI), defined as focal ischemic damage on conventional magnetic resonance imaging (MRI) without clinical signs or symptoms of stroke (4–6). SCI is an independent risk factor for overt stroke and is associated with lower scores on math and reading achievement tests, Full-Scale Intelligence Quotient (FSIQ), Verbal IQ, and Performance IQ (5–7).

In patients with SCD, neurocognitive deficits arise during early childhood and persist into adulthood, negatively affecting their health outcomes and quality of life, as well as increasing disease complications (8). Neurocognitive deficits are also exacerbated by environmental factors, such as low socioeconomic status (9). In a meta-analysis of 110 studies involving 3600 participants with SCD, deficits in FSIQ, verbal reasoning, perceptual reasoning, and executive function increased from preschool- to school-aged participants (Prussien et al., 2019). Furthermore, more severe anemia, lower total Hb and fetal Hb, cerebral occlusion, and reduced oxygen saturation are all associated with more severely impaired cognitive processing in SCD (11). In some cases, however, neurocognitive deficits occur without neurologic injury, i.e., in individuals with normal diagnostic imaging (Prussien et al., 2019; Schatz, 2002).

The current widely used standard treatment for SCD is hydroxyurea. Patients receive daily oral administration of hydroxyurea, often throughout their lifetimes. Several studies have attempted to identify the effects of hydroxyurea treatment on neurocognitive performance in children with SCD (14–17). Among these, Puffer et al. (2007) reported an improved global cognitive index with hydroxyurea treatment, and Heitzer et al. (2021) found an association of hydroxyurea therapy with higher neurocognitive measures across multiple assessments. Wang et al. (2021) showed significantly improved reading comprehension after hydroxyurea treatment in patients with SCD. However, there is limited clinical neuroimaging data relevant to hydroxyurea therapy.

Understanding the neural correlates of deficits in SCD, the treatment effects of hydroxyurea on brain function, and the associated mechanisms is important. Task-based fMRI approaches have been developed to investigate the brain regions related to neurocognitive deficits in SCD. Among neurocognitive deficits, working memory is a critical area of impairment because it has a significant impact on quality-of-life outcomes, including academic development and achievement (18,19). Furthermore, working memory is commonly affected in youth with SCD (20). Brain regions with working memory functions are relatively more vulnerable to SCD. For instance, disrupted oxygen delivery to deep white matter, basal ganglia, middle and superior frontal gyrus, and dorsal parietal regions are often found in SCD patients due to derangement in cortical and subcortical structures at the distal portions of the anterior and middle cerebral artery (1,21). To assess working memory, the n-back test is widely used in cognitive neuroscience (22). By using a computerized visual–spatial n-back task, some studies have shown evidence of poorer visual–spatial working memory in children with SCD, compared to demographically matched healthy controls (20,23). To identify the brain regions involved in working memory processing and its deficits in patients with SCD and to trace the longitudinal neurocognitive effects of hydroxyurea treatment, task-based functional magnetic resonance imaging (fMRI) at baseline and follow-up time points is necessary.

In this study, we hypothesized that hydroxyurea improves working memory function and impacts working memory processing in the brain of patients with SCD. We also aimed to identify hydroxyurea treatment effects in brain regions related to working memory processing. To test our hypothesis, fMRI data during n-back tasks using images of objects as stimuli were acquired in pediatric patients with SCD. The fMRI sessions were performed at two time points, before hydroxyurea treatment, and at 1-year follow-up. Neurocognitive measures were also assessed separately at both time points. We also measured the longitudinal effects of hydroxyurea treatment in behavior performance of the n-back tasks and neurocognitive tests. Subsequently, we analyzed brain-activation patterns during working memory processing and identified brain regions with changes in activation at 1-year follow-up via searchlight-pattern classification. Next, we analyzed changes in BOLD signals during working memory processing as a function of time points or working memory load. We also investigated if changes in BOLD signals during working memory processing were correlated to changes in behavior performance of the 2-back task across time points.

## Method

### 2.1 Participants and treatment

Patients (7–18 years old) who had a diagnosis of SCD (HbSS or HbSβ^0^-thalassemia) and had not been previously exposed to hydroxyurea but met clinical criteria to receive this therapy were recruited to participate in this study. Participants were excluded from participation if they had a history of clinical stroke, were receiving chronic transfusion therapy, had received a hematopoietic stem cell transplant, or could not tolerate MRI examination without sedation or anesthesia. Participants were recruited from the Sickle Cell Outpatient Clinic at St. Jude Children’s Research Hospital (St. Jude) between June 2011 and January 2015, when they were initiating hydroxyurea therapy. This study was approved by St. Jude IRB. Written informed consent was obtained from each participant or from the legal guardian of each minor participant. Each minor participant also gave verbal assent before participating in the research exams.

A total of thirty-one patients with SCD were approached and consented to the study. Eleven participants were assigned to the control group because they never initiated hydroxyurea treatment or discontinued the drug shortly after starting it. These participants had a mean age of 11.9 years (SD 2.5 years, range 7.6–16.5 years) at enrollment; 3 (27%) were female. The 20 patients who received hydroxyurea therapy continuously were assigned to the treated group. These participants had a mean age of 12.3 years (SD 3.3 years, range 7.1–17.3 years) at enrollment; 12 (60%) were female.

### 2.2 Study design

The study protocol was approved by the St. Jude Institutional Review Board. A parent or legal guardian of each participant gave informed consent, and participants gave assent, as appropriate. Participants were evaluated before starting hydroxyurea therapy. Baseline MR imaging, transcranial Doppler ultrasound examination, neurocognitive testing, and blood work were completed, usually over 2 days. In the treated group, hydroxyurea dosing was initiated at 20 mg/kg/day and gradually escalated per the standard protocol (24). All evaluations were conducted at 2 time points, baseline before treatment and 1-year follow-up, and compared between 2 groups: participants who received hydroxyurea treatment (HU group) and those who were not (non-HU control group).

### 2.3 Magnetic resonance imaging

Siemens MRI 3T scanners were used in this study. A 3D T1-weighted sagittal MPRAGE sequence (TR/TE 1980 ms/ 2.26 ms, 1.0-mm pixel space, 256 × 256 matrix, 160 slices, and 1.0-mm slice thickness) was acquired to facilitate spatial processing of functional images and visualize fMRI results. The fMRIs were performed with a T2* weighted echo-planar imaging sequence with the following parameters: TR =2.0 s, TE = 30 ms, flip angle = 90°, field-of-view = 192 mm x192 mm, matrix = 64 x 64, slice thickness = 3.5 mm, and total 32 slices in one volume. Participants were trained for the fMRI scanning sessions with computerized programs before entering the scanner room. The head was immobilized with vacuum pads in the head coil during the scan sessions, and fMRI instructions and stimuli were viewed through the head coil mirror.

### 2.4 Object n-back tasks

Participants were tested with 0-, 1-, and 2-back tasks using objects (abstract images not easily namable, Fig. 1) as stimuli while undergoing fMRI scanning. Each n-back task was repeated 3 times with the order of 0-, 1-, and 2-back (i.e., total = 9 blocks). For the 0-back task, participants responded by pressing a button to detect a specified target, which is the first image in Fig. 1. For the 1- and 2-back tasks, participants responded if the current image was identical to the previous image (1-back) or the image presented 2 trials earlier (2-back). Before each task, instructions associated with each n-back task were displayed for 2 s, followed by 5 s of rest. During each task block, a series of 16 stimuli, including 4 targets, was presented. Each stimulus was displayed for 500 ms, with a 1500 ms interval between stimuli. Reaction time, omissions, and commissions were recorded during all n-back tasks. In addition, balanced accuracy (BA), which is the arithmetic mean of sensitivity and specificity, was measured using the equation shown below. BA was used instead of “overall” accuracy because the total numbers of targets (n = 12) and nontargets (n = 36) were imbalanced.

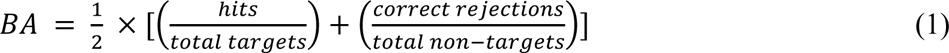

**Figure 1.**
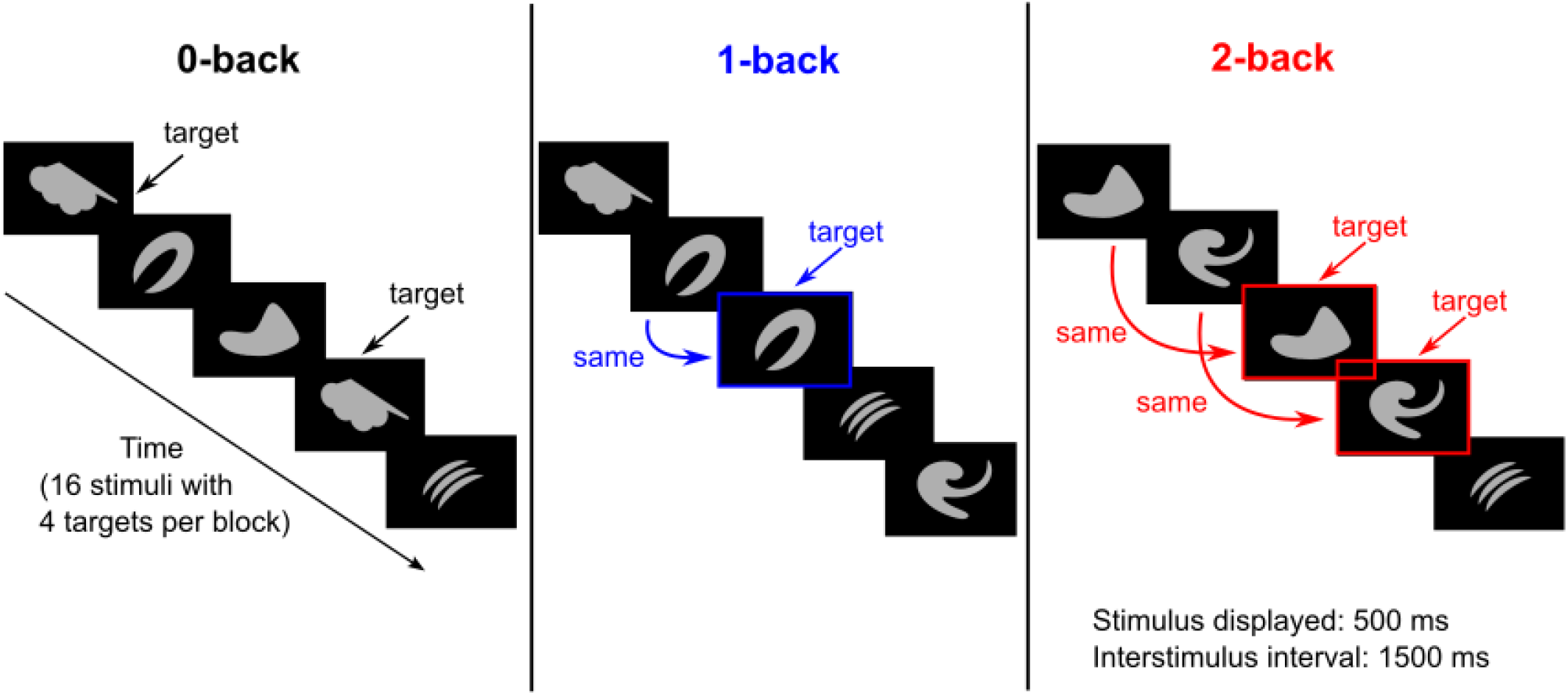
Schematic overview of the n-back task using images of objects as stimuli. The 0-, 1-, and 2-back tasks were performed separately in each block.

### 2.5 Neurocognitive tests

Neurocognitive measures sensitive to the deficits in SCD were chosen and tested. Measures were chosen with appropriate test-retest reliability and negligible practice effects for the 1-year interval between testing time points. The intellectual function of participants was tested with the Wechsler Intelligence Scale for Children, Fourth Edition (WISC-IV; Weschler D., 2003). The WISC was used to assess intelligence in children 7–17 years of age, and the Wechsler Adult Intelligence Scale, Fourth Edition (WAIS-IV; Weschler D., 2008) was used for participants 18 years and older. A shortened administration of 8 subtests allowed the derivation of the index scores of FSIQ, verbal comprehension, perceptual reasoning, working memory, and processing speed.

### 2.6 fMRI pre-processing and analysis

The fMRIs were pre-processed and analyzed with SPM software (www.fil.ion.ucl.ac.uk/spm). For first-level analyses, SPM 8 was used; for group-level analyses, SPM 12 was used. Motion-corrected fMRI time series from the MRI scanner were used in fMRI image analysis. Pre-processing included slice timing correction, realignment, co-registration to the 3D-T1 image, spatial normalization to the Montreal Neurological Institute brain template, and spatial smoothing with an 8 mm^3^ FWHM Gaussian kernel. Individual contrast maps from the first-level analysis were used in a full-factorial second-level analysis model to assess the group effects, time point effects, and group and time interaction effects.

### 2.7 Searchlight analysis

The details of searchlight analysis were presented by Kriegeskorte et al. (2006). In this method, the imaged volume of the brain is scanned with a “searchlight,” and the contents undergo multivariate classification at each location in the brain. In brief, a 3D sphere with a given radius (usually in millimeters) is scanned across the whole brain or regions of interest. A classifier or regressor is then trained on the corresponding voxels to predict the experimental, relevant behavior or condition, and prediction accuracy is measured for each corresponding voxel.

We performed searchlight analysis with a software program developed in-house in the Python language using the ‘nilearn’ package (28). During the analysis, we first isolated object-evoked fMRI signals during the 2-back task measured at baseline and 1-year follow-up from participants with fMRI data at these 2 time points. At each time point, BOLD signals of 2-back tasks were obtained by subtracting the average 0-back object-evoked fMRI signals from each trial of 2-back object-evoked fMRI signals. We retrieved 42 trials of BOLD signals in the 2-back task at each time point. Next, we performed searchlight analysis over the whole brain to identify the areas containing information to classify BOLD signals into those measured at pre-treatment vs. 1-year follow-up. The same searchlight analysis was conducted separately in the treated (13 participants) and the control (8 participants) groups. We used a sphere radius of 4 mm, leave-one-participant-out cross validation, and a k-nearest neighbor classifier (29). According to Kriegeskorte et al. (2006) and Etzel et al. (2013), a 4-mm radius usually yields near-optimal detection performance. After testing different classifiers, we concluded that the k-nearest neighbor classifier provided the best classification accuracy. Neural discrimination accuracy using this classifier was also reported to be highly correlated with animal behavior (31,32).

Fig. 2 illustrates a complete process of searchlight analysis within the treated or the control group. The searchlight analysis produced an accuracy score (0 to 1.0) for each voxel in which 0 indicates no predictive power and 1.0 indicates best predictive power. To identify specific brain regions that contained information with higher accuracies to distinguish BOLD signals measured at pre-treatment vs. 1-year follow-up, we applied a Python package named ‘AtlasReader’ (33) to identify significant clusters and the anatomical locations of these clusters. A cut-off threshold for an accuracy score of 0.65, a threshold for a cluster size of 20, and the AAL atlas (34) were used. Among all the clusters derived from searchlight analyses in the treated and control groups, we selected clusters with larger volumes (≥ 520 mm^3^) and higher peak accuracies (≥ 0.7) for subsequent analyses. The median of BOLD signals in each selected cluster at each time point and during different levels of n-back tasks were computed for comparisons. The BOLD signal differences between time points in each voxel of each selected cluster were also computed for subsequent analyses of brain-behavior relationships.

**Figure 2.**
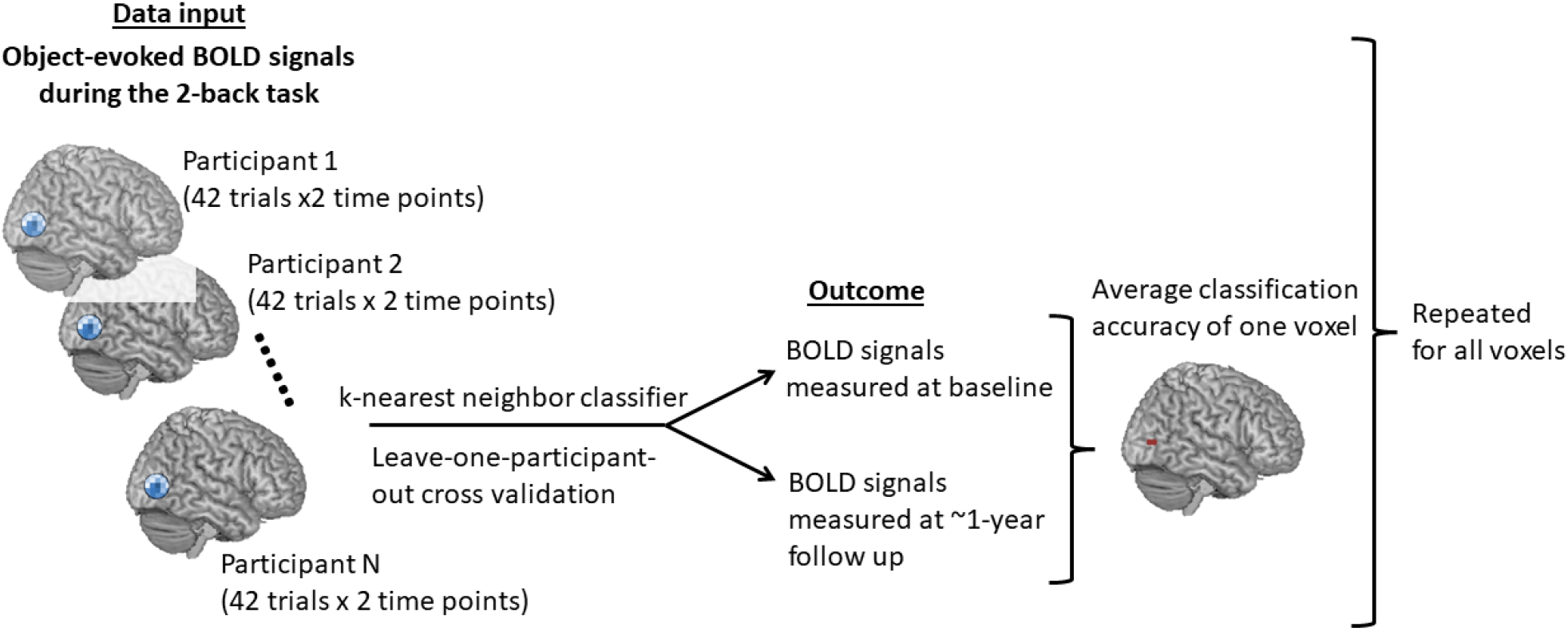
Schematic overview of the searchlight-pattern classification conducted in the HU and non-HU groups separately.

### 2.8 Statistical analyses

We compared n-back task performance (including reaction time, BA, commission errors, and omission errors), neurocognitive measures, and the medians of 2-back BOLD signals for each selected cluster across time points (within group) or across groups (between groups). For n-back task performance and the medians of 2-back BOLD signals, the Wilcoxon rank-sum test was used to compare variables between the 2 groups at a single time point. Meanwhile, the Wilcoxon signed-rank test was used to compare variables between the 2 time points within the same group. For neurocognitive measures, the Student’s *t*-test was used for between-group comparison at a single time point while the paired sample *t*-test was used for within-group comparison across time points. In addition, we compared the medians of BOLD signals for each selected cluster at a 1-year follow-up time point as the working memory load increased from 0- to 2-back. The Wilcoxon signed-rank test was used to compare BOLD signals between task difficulties within the same group. All these analyses were exploratory in nature due to the limited number of observations in each condition and as a result, we did not adjust analyses for multiple comparisons. For these analyses, *p*-values were 2-sided, and *p* <.05 was considered statistically significant.

### 2.9 Brain and behavior relationships

We explored the association between 2-back BOLD signal differences (i.e., data of 1-year follow-up subtracted data of pre-treatment) and behavior performance differences of the 2-back task for each selected cluster. Since a significant increase in omission errors in the 2-back task was found in the non-HU group across time points (Fig. 3D), we focused on investigating the omission error change and BOLD signal change between two time points for the 2-back task. Only participants with data on omissions errors and BOLD signals at both time points were used in this analysis. We fitted a regression line (eq. 2) with BOLD signal differences (*BOLD_diff*), omission error differences (*OM_diff*), and group (*grp*: non-HU control = 0 & HU treated = 1) for each of the selected clusters separately.

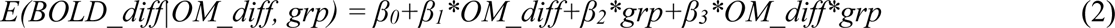

**Figure 3.**
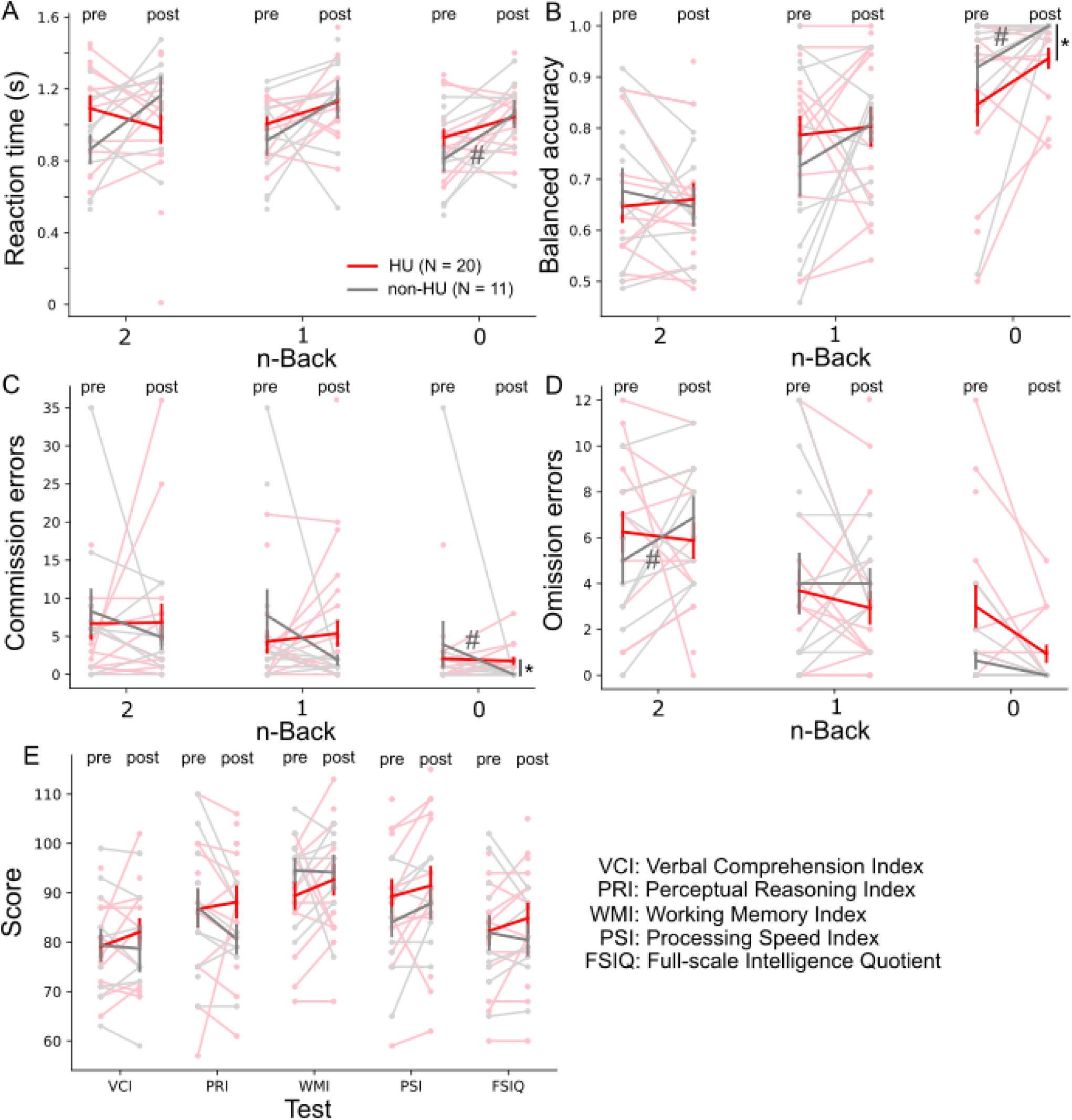
Hydroxyurea (HU) treatment did not significantly affect performance on the 0- to 2-back tasks or on the neurocognitive tests. Reaction time (**A**), balanced accuracy (**B**), commission errors (**C**), and omission errors (**D**) of 0-,1-, and 2-back tasks were measured pre-treatment and 1-year follow-up. Neurocognitive tests (**E**) were also measured at pre-treatment and 1-year follow-up (post). When compared across HU and non-HU control groups at the 1-year follow-up time point, the balanced accuracy and commission errors of the 0-back task were significantly different. Red and gray bold lines represent the means of the HU and the non-HU control groups, respectively. Light red and light gray markers represent individual data points in the HU and the non-HU control groups, respectively. * *p* <.05 (Wilcoxon rank-sum test) and # *p* <.05 (Wilcoxon signed-rank test).

In the above equation, *β_0_* is the intercept, *β_1,_* and *β_2_* are the coefficients for differences in behavior performance between time points and group respectively while *β_3_* is the coefficient for the interaction of behavior differences and group. We focused on the significance of the slopes (*β1* for the control group and *(β1+ β3)* for the treated group) of the fitted regression lines for each selected cluster in each group. The slopes (*β1* and *β1+ β3*) indicate the change in *BOLD_diff* with a unit change in *OM_diff* in the control group and the treated group, respectively.

## Results

### 3.1 Hydroxyurea treatment

For the HU group, the initial hydroxyurea dose was 20 mg/kg/day, and doses were gradually adjusted to an average of 23.8 mg/kg/day (range 13.3–33.4 mg/kg/day) after 1 year of treatment. Hematologic responses to hydroxyurea at 1 year suggested good adherence to treatment (17). The Hb level, fetal Hb level, absolute reticulocyte count, and other measures have been previously reported (Wang et al., 2021).

### 3.2 Performance of n-back tasks and neurocognitive measures

Although all participants completed the n-back (0-,1-, and 2-back) tasks while simultaneously undergoing fMRI scanning at both time points, some participants’ data at one or both time points were excluded due to imaging quality concerns. For the HU group, 6 participants were excluded in the pre-treatment time point and 4 participants were excluded in the 1-year follow-up time point. In the non-HU control group, 3 participants were excluded in the 1-year follow-up time point. In Fig. 3A-D, we did not observe a significant treatment effect within the HU group (Wilcoxon signed-ranked test, *p* > .05) when all n-back tasks were compared across time points. Within the non-HU control group, significant differences were observed in the reaction time, BA, and commission errors in the 0-back task when compared across time points. The omission errors were found to increase significantly in the 2-back task when compared across time points within the non-HU control group. At the 1-year follow-up time point, we did not observe differences between the HU and non-HU control groups (Wilcoxon rank-sum test, *p* > .05), except in the BA and commission errors of the 0-back task. Although there were more participants with the neurocognitive measures, we did not observe statistically significant differences between the 2 groups (Student’s *t*-test, *p* > .05) or the two time points (paired sample *t*-test, *p* > .05) (Fig. 3E).

### 3.3 fMRI interaction effect of the treatment group and time point

We tested various contrasts including 1-> 0-back, 2-> 1-back, 2-> 0-back, 2-back > resting-condition, etc., using full factorial modeling with both groups and time points to search for main and interaction effects. We also compared, for example, 2-> 0-back contrast between pre-treatment and ∼1-year follow-up in the HU and non-HU control groups separately. Among all the tests and comparisons, we detected a significant interaction effect (T_48_ = 3.27, *p* = .0001) of the treatment group and time point in the right cuneus and angular gyrus only for the 2-> 0-back contrast in participants from both groups (Fig. 4). The main and interaction effects of the treatment group and time point were not significant when we used the other contrasts.

**Figure 4.**
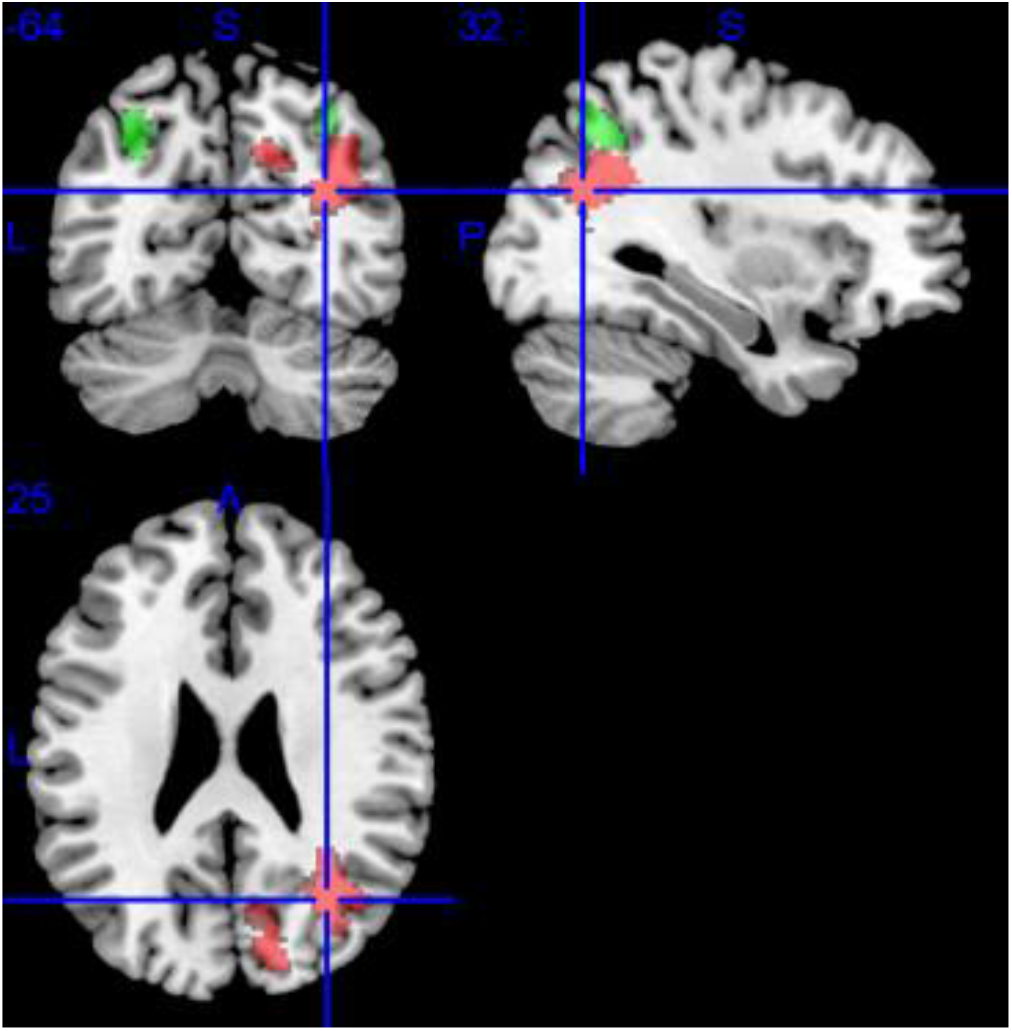
A significant treatment-by-time interaction was detected in the right cuneus and angular gyrus (labeled in red) for 2-> 0-back contrast. The brain regions activated by the n-back tasks using objects as stimuli are labeled green. Blue lines marked the MNI coordinate [32, −64, 25].

### 3.4 Brain regions of searchlight-pattern classification

We did not observe significant treatment effects with hydroxyurea when performing traditional fMRI univariate analysis; thus, we shifted our analytical strategy to multivariate pattern analysis. Searchlight-pattern classification is a type of multivariate classification at each location in the brain that helps identify regions predictive of experimental conditions (27). We performed searchlight analyses to identify and localize brain regions or clusters that predicted 2-back BOLD signals recorded at pre-treatment or 1-year follow-up (see Fig 2). These analyses were conducted separately in the HU and the non-HU groups. Brain regions or clusters with higher classification accuracy scores reported in the results were those with more prominent changes in brain activation between time points during the 2-back task. Clusters with classification accuracies > .65 and size > 20 identified from each group are colored in red and shown in Fig. 5. A total of 27 clusters, with most larger clusters localizing in the posterior region, were reported in the non-HU group. On the other hand, 7 clusters, with only 2 larger clusters localizing in the frontal region, were reported in the HU group. Next, we selected clusters with peak accuracy ≥ .7 and volume ≥ 520 mm^3^ for downstream analyses. Among all the identified clusters in both groups, 7 clusters in the non-HU group and 2 clusters in the HU group fulfilled the selection criteria. The anatomical locations and the main regions in the brain of these 9 selected clusters are shown in Fig. 6. Moreover, the details of these clusters, including peak MNI coordinates, volume, brain regions with % of coverage in AAL, etc., are reported in Tables 1 (non-HU group) and 2 (HU group).

**Figure 5.**
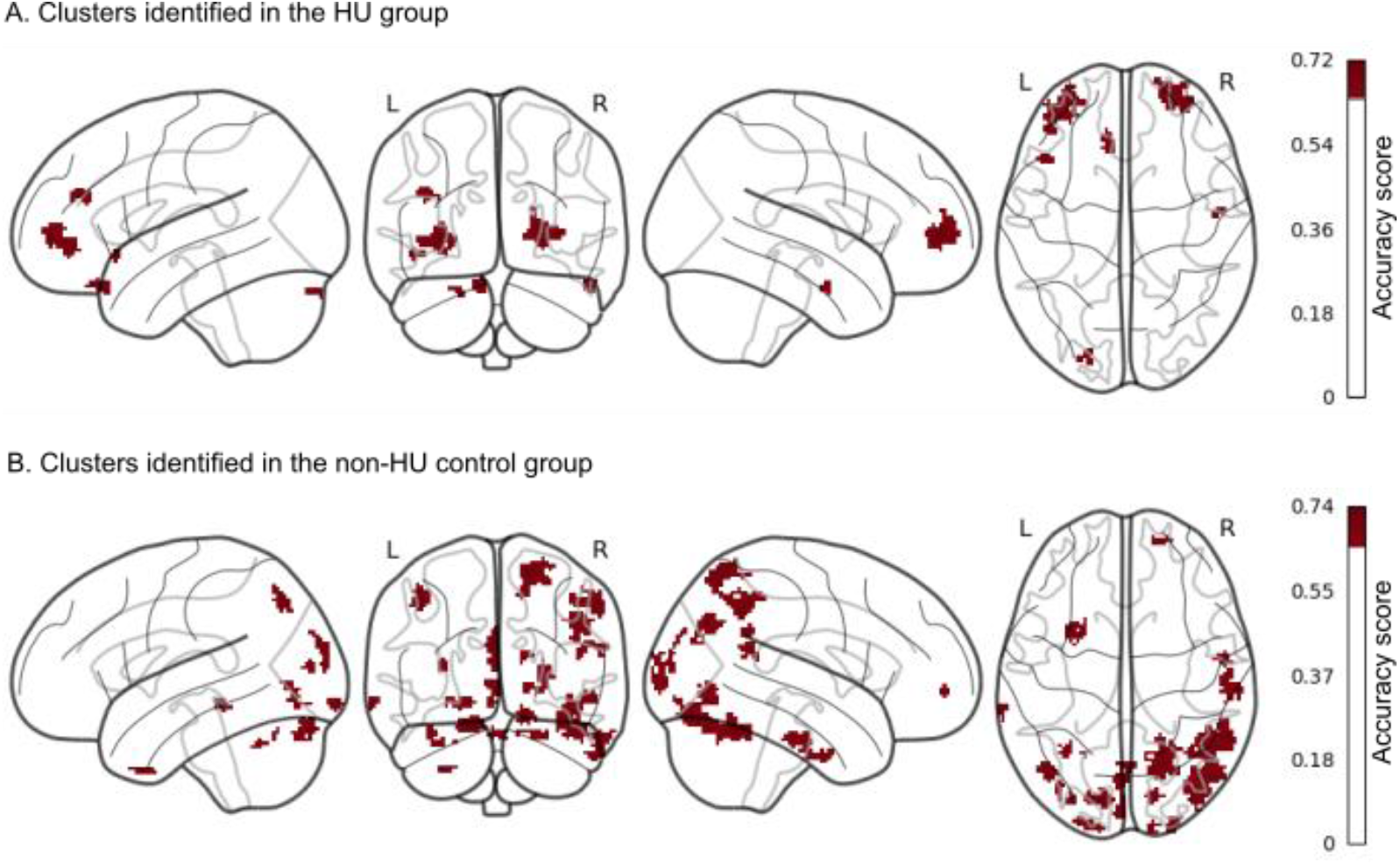
More clusters with changes in brain activation between time points during the 2-back task were identified in the non-HU control group than in the HU group. Clusters with higher predictive power of time points were identified using a classification accuracy threshold of > .65 and a cluster-size threshold of > 20. These clusters had larger differences in 2-back BOLD signals between time points. In the non-HU group, 27 clusters, with most larger clusters localizing in the posterior region, were identified. In the HU group, 7 clusters with only 2 larger clusters localizing in the frontal region were reported.

**Figure 6.**
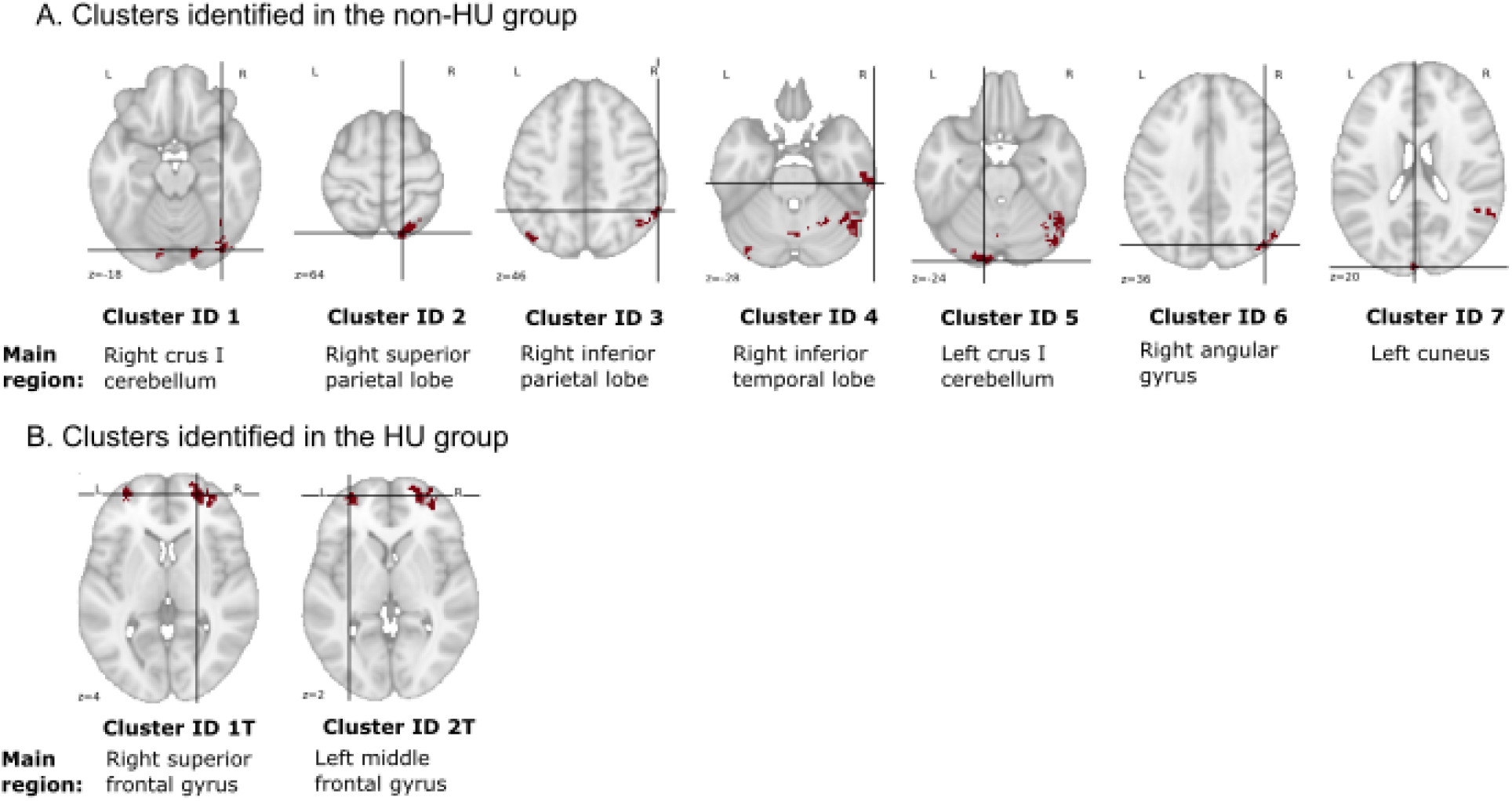
Anatomical locations of the 9 selected clusters (with peak value ≥ .7 and volume ≥ 520 mm^3^) at the transverse plane and their respective main regions in the brain (the highest percentage % coverage in AAL). The details of these clusters are reported in Tables 1 and 2. The crossed lines marked the peaks of the clusters.

**Table 1.**
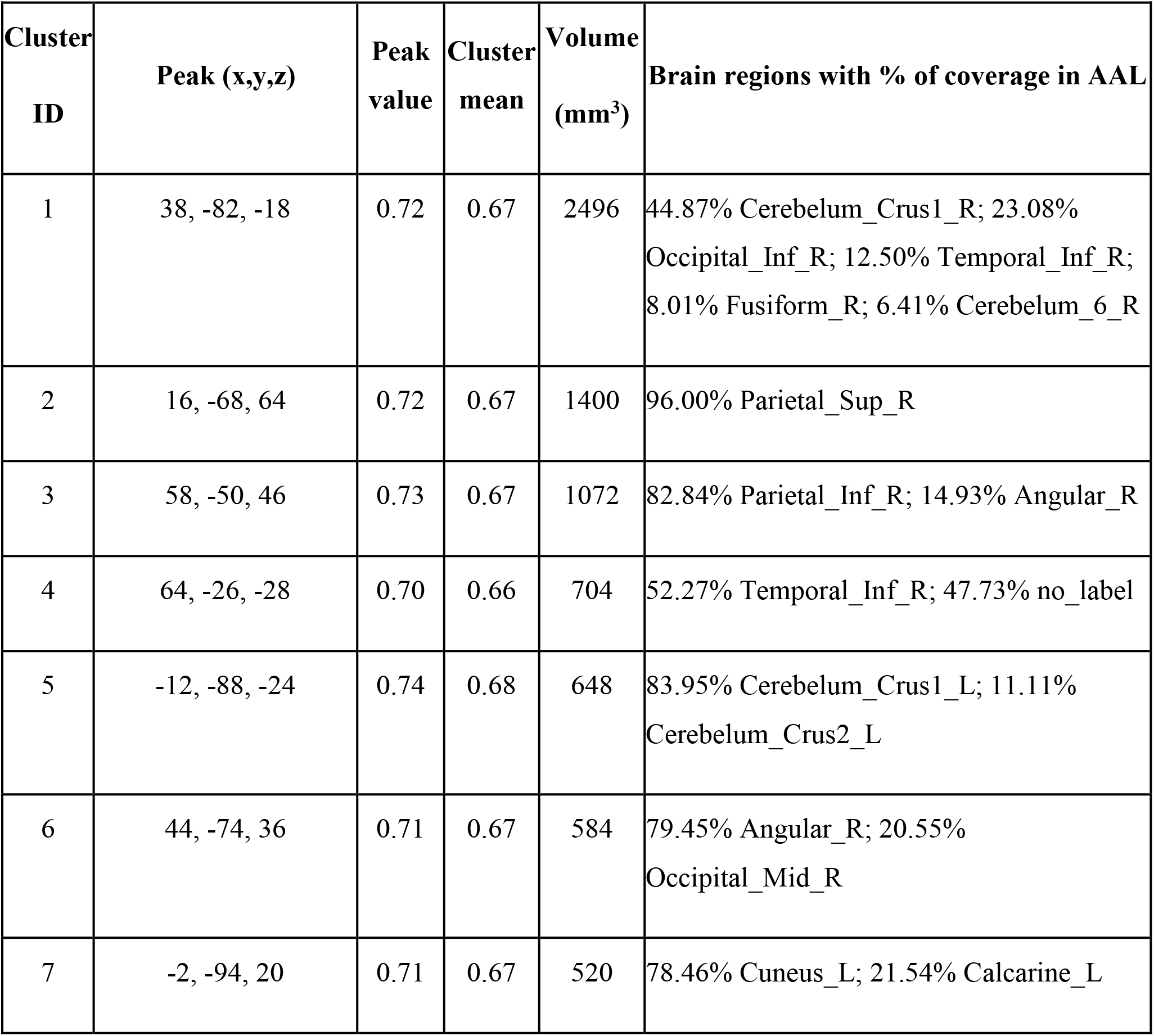
Clusters (peak accuracy ≥ .7 & volume ≥ 520 mm^3^) identified in the non-HU group.

**Table 2.** Clusters (peak accuracy ≥ .7 & volume ≥ 520 mm^3^) identified in the HU group.

| Cluster ID | Peak (x, y, z) | Peak value | Cluster mean | Volume (mm<sup>3</sup>) | Brain regions with % of coverage in AAL |
| --- | --- | --- | --- | --- | --- |
| 1T | 24, 56, 4 | 0.72 | 0.67 | 1744 | 75.69% Frontal_Sup_2_R; 24.31% Frontal_Mid_2_R |
| 2T | -32, 58, 2 | 0.71 | 0.66 | 1368 | 53.80% Frontal_Mid_2_L; 44.44% Frontal_Sup_2_L |

### 3.5 Brain activation during working memory processing

For each of the 9 selected clusters, we computed the median of 2-back BOLD signals at the 2 time points, respectively, in both groups. We also computed the median of BOLD signals for each selected cluster during 0-, 1- or 2-back tasks at 1-year follow-up. We observed increases in the medians of 2-back BOLD signals in most of the selected clusters at 1-year follow-up (post) compared to the baseline (Fig. 7A). In the non-HU control group, 6 of the selected clusters (corresponding to the right crus I cerebellum, right inferior parietal lobe, right inferior temporal lobe, right angular gyrus, left cuneus and left middle frontal gyrus) had higher BOLD signals during working memory processing at 1-year follow-up (*p* < .05, Wilcoxon signed-rank test). However, only 1 of the selected clusters (the right superior parietal lobe) had higher BOLD signals in the HU group at 1-year follow-up (*p* < .05, Wilcoxon signed-rank test). In addition, when changes in BOLD signals were analyzed as a function of task difficulty at 1-year follow-up (Fig. 7B), increases in BOLD signals (*p* < .05, Wilcoxon signed-rank test) were observed when the task difficulty increased from 0- to 1-back in most selected clusters in both groups. In the non-HU control group, BOLD signals elevated as the working memory load increased from 0- to 1-back, but not from 1- to 2-back, in the right inferior temporal lobe, right angular gyrus, and right superior frontal gyrus. However, BOLD signals continued to increase (*p* < .05, Wilcoxon signed-rank test) in the left crus I cerebellum, right angular gyrus, left cuneus and right superior frontal gyrus of the HU group as the working memory load increased from 0- to 2-back.

**Figure 7.**
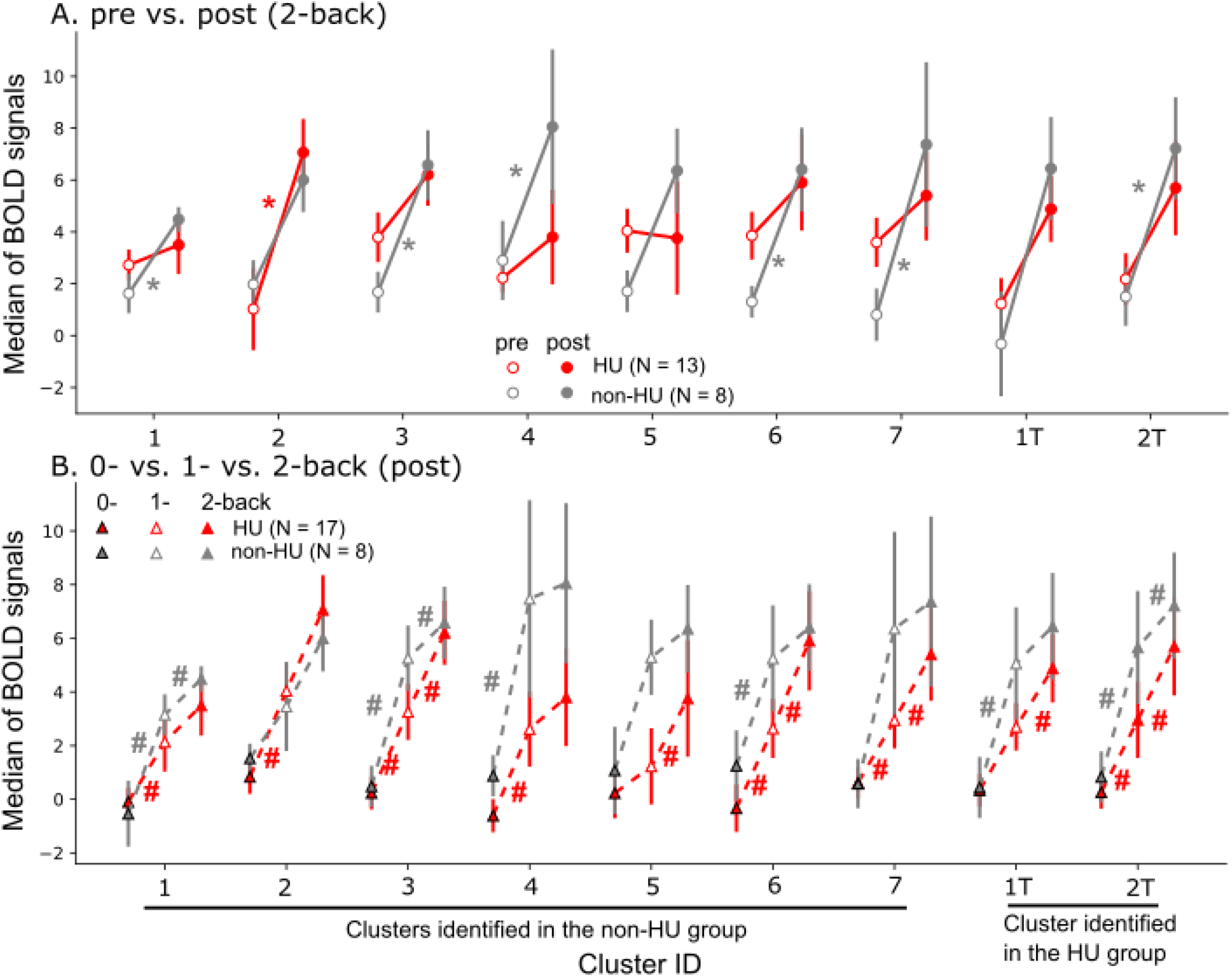
Increased BOLD signals during working memory processing as a function of time points and task difficulty. Nine clusters (refer to Fig. 6, Tables 1 & 2) were selected from all the clusters identified via searchlight analyses. (A) Six selected clusters in the non-HU control group had higher brain activation during working memory processing at 1-year follow-up (post) compared to pre-treatment. However, only one selected cluster had higher brain activation in the HU group at 1-year follow-up compared to pre-treatment. * *p* < .05 [Wilcoxon signed-rank test within the HU group (red) or the non-HU group (gray)]. (B) Increased BOLD signals were observed in most selected clusters in both groups as the working memory load increased from 0- to 1-back. In the non-HU control group, BOLD signals did not increase further when the working memory load increased from 1- to 2-back. In the HU group, however, BOLD signals increased continually as the working memory load increased from 0- to 2-back. # *p* < .05 (Wilcoxon signed-rank test).

We analyzed the relations of differences in 2-back BOLD signals and differences in omission errors between time points for each selected cluster. These analyses were exploratory in nature as we had a limited number of participants (N = 8 in the non-HU group & N = 12 in the HU group) with fMRI data and omission errors at both time points. Among all the fitted regression lines (*BOLD_diff* vs. *OM_diff*), we did not observe statistical significances in the slopes in either group, which indicates we did not find the difference of the BOLD signal between two time points was associated with the omission error changes between the same time points in both groups.

## 4 Discussion

In this study, we evaluated the longitudinal neurocognitive effects associated with hydroxyurea treatment and identified changes in brain activation during working memory processing in specific brain regions in patients with SCD. We did not observe any significant hydroxyurea treatment effect in behavior measures of all n-back tasks and fMRI group-level analyses of various contrasts. However, in the fMRI group-level analysis of 2-> 0-back contrast, we detected increased BOLD signals in the HU group but reduced BOLD signals in the non-HU control group in the right cuneus and angular gyrus when compared between baseline and 1-year follow-up time points. However, when we conducted searchlight-pattern analyses to classify 2-back BOLD signals into those measured at baseline or 1-year follow-up, we identified more changes in the pattern and magnitude of BOLD signals in the posterior part of the brain across time points in the non-HU group than in the HU group. Elevated 2-back BOLD signals were observed in the right crus I cerebellum, right inferior parietal lobe, right inferior temporal lobe, right angular gyrus, left cuneus and left middle frontal gyrus at 1-year follow-up in the non-HU group. Increased 2-back BOLD signals were only found in the right superior parietal lobe in the HU group at 1-year follow-up. When the demand for working memory increased, BOLD signals in the non-HU group elevated from 0- to 1-back but did not increase further from 1- to 2-back at 1-year follow-up in the right inferior temporal lobe, right angular gyrus, and right superior frontal gyrus. However, BOLD signals in the HU group increased with increasing working memory load from 0- to 2-back at 1-year follow-up in the left crus I cerebellum, right angular gyrus, left cuneus and right superior frontal gyrus.

Surprisingly, our findings revealed more changes in the pattern of brain activation and increased BOLD signals during working memory processing with no hydroxyurea treatment in the non-HU control group at 1-year follow-up. In contrast, fewer changes in the pattern and magnitude of brain activation during working memory processing with hydroxyurea treatment in the HU group were observed at 1-year follow-up.

### 4.1 Effect of hydroxyurea treatment on cognition

Hydroxyurea treatment is a standard of care for children with SCD, and several studies have shown positive effects of hydroxyurea treatment on neurocognitive functions. One study reported improved verbal comprehension, global cognitive ability, and fluid reasoning in 15 patients with SCD (aged 6–21 years) after at least 1 year of hydroxyurea treatment compared to 50 untreated patients (14). Our recent study showed a notable trend of improvement in FSIQ and a significant gain in reading comprehension in 19 patients with SCD (aged 7.2–17.8 years), comparing before and after 1 year of hydroxyurea treatment (17). Moreover, another cross-sectional study assessing neurocognitive performance in 364 patients with SCD (aged 8–24 years) of multiple age groups revealed that early treatment with hydroxyurea potentially reduces, but does not eliminate, neurocognitive decline with age (15).

The mechanism of how hydroxyurea improves cognitive outcomes is not completely clear (14). One potential mechanism is that hydroxyurea suppresses cell stress signaling, leading to increased levels of fetal hemoglobin and hemoglobin (35) that cause positive effects on brain oxygenation and function. As hydroxyurea decreases anemia and thrombocytosis (36,37), it may prevent or reverse functional brain abnormalities, slowing down the process of neurocognitive declines. In addition to impacting hemoglobin level and brain oxygenation, hydroxyurea may lead to improved cognitive performance more indirectly by decreasing general illness severity and pain (38).

In this study, hydroxyurea treatment did not affect behavioral measures of reaction time, BA, omission errors, or commission errors, nor did it affect any of the neurocognitive measures as assessed by fMRI. However, in patients receiving no hydroxyurea treatment, increased omission errors in the 2-back task were observed when compared across time points within the group. Moreover, changes in reaction time, balanced accuracy and commission errors were found within the non-HU group when compared across time points. The lack of significant improvements with hydroxyurea treatment in other neurocognitive measures and the n-back tasks could be because a longer time interval (i.e., more than 1 year) between assessments may be required to detect significant changes in behavior performance of working memory and neurocognitive tests. Furthermore, the neurocognitive effects of hydroxyurea may depend on when patients start the treatment. Hydroxyurea may produce greater neurocognitive effects in patients who start the treatment earlier in life, during the critical period of development, or in those with a lower baseline ability (17).

Although we observed no changes in most behavior measures of the 2-back task, changes in brain activation during the 2-back task were identified via searchlight-pattern analyses in the non-HU and HU groups. Searchlight-pattern classification of 2-back BOLD signals into those measured at baseline vs. 1-year follow-up reported more clusters with higher accuracy scores (> .65) in the non-HU group than the HU group. More voxels with higher classification accuracies were obtained when there were more changes in brain activation across time points. In contrast, fewer changes or more similarities in brain activation across time points reduced classification accuracies. As a result, these findings suggested that patients with SCD with hydroxyurea treatment had less changes in brain activation during working memory processing. This phenomenon could be an effect of hydroxyurea treatment in maintaining working memory function from degrading over time in patients with SCD. Therefore, changes in behavior performance in the 2-back task with hydroxyurea treatment were not noticed at 1-year follow-up.

### 4.2 Increased brain activation during working memory processing

In addition to identifying more changes in brain activation during working memory processing in the non-HU group at 1-year follow-up, we observed increased brain activation in 6 (the right crus I cerebellum, right inferior parietal lobe, right inferior temporal lobe, right angular gyrus, left cuneus and left middle frontal gyrus) out of the 9 selected clusters in patients with no hydroxyurea treatment but only 1 cluster (the right superior parietal lobe) with increased brain activation in the HU group at 1-year follow-up. Larger increases in BOLD signals were observed in the non-HU group as the working memory load increased from 0- to 1-back compared to from 1- to 2-back. These observations in the non-HU group suggested an increase in cognitive effort during working memory processing in patients with SCD with no hydroxyurea treatment at 1-year follow-up and as working memory load increased from 0- to 1-back. An elevation in brain activation in the salience network as a function of increasing demands of working memory tasks was reported by Engström et al., (2013). They observed a steeper increase of the BOLD signal as a function of increasing cognitive effort in participants with low working memory capacity. While BOLD signals in the non-HU group did not increase further as the working memory load increased from 1- to 2-back, BOLD signals increased continually with increasing working memory load from 0- to 2-back in the HU group. This coincides with the study of Engström et al., (2013) in which high working memory performers were found to have BOLD responses modulated by effort along a larger dynamic range.

In addition, some of the selected clusters (refer to Tables 1 & 2) have main brain regions, such as the inferior parietal lobe, right Crus I cerebellum, and right angular gyrus, that are related to attention, working memory and effortful processing. For example, cluster ID 1 was located partially at the right Crus I of the cerebellum and right inferior occipital lobe. The Crus I of the cerebellum was reported to contribute to working memory in previous studies (40,41) while the occipital lobe has a role in object recognition and memory formation (42). Cluster ID 3 was in the right inferior parietal lobe and the right angular gyrus. Both brain regions are involved in spatial attention (43) and memory (44). Cluster ID 1T was in the right superior frontal gyrus and middle frontal gyrus. The superior frontal gyrus is a key component in the neural network of working memory (45) while the middle frontal gyrus has a role in reorienting attention (46). Therefore, in this study, we found increased BOLD signals at 1-year follow-up in brain regions associated with attention, working memory and effortful processing potentially due to cognitive degradation with no hydroxyurea treatment in patients with SCD.

### 4.4 Limitations and future study

One of the limitations of this study is the small number of participants with complete fMRI and behavior data of n-back tasks at both time points. Moreover, the 2-back task is more difficult than the 1- and 0-back tasks; thus, some participants had problems understanding the task and failed to perform it per instructions. The participants’ baseline working memory and some neurocognitive measures before treatment were slightly different and might confound the results. Baseline measurements were obtained at various ages because participants started hydroxyurea treatment at different ages; the severity of SCD also differed among the participants. We worked around these limitations by conducting multivariate pattern analysis, which has higher sensitivity, even with smaller sample sizes. Although traditional fMRI univariate analysis has a limited ability to identify subthreshold brain signals, multivariate pattern analysis gets around that limitation by analyzing brain signals in a multidimensional space, which has the potential for novel discoveries (47).

Hydroxyurea is a disease-modifying treatment, and patients need to take the drug throughout their lifetime for continued effect. Therefore, a longitudinal study that evaluates neurocognitive changes over a longer period is essential for obtaining more conclusive outcomes. Furthermore, curative treatments, like gene therapy (48) and hematopoietic cell transplantation (49) may produce more positive neurocognitive effects than hydroxyurea does after treatment. Therefore, future studies may investigate brain activation patterns and neurocognitive changes during n-back tasks and other cognitive tasks in patients who receive curative treatments.

### 4.5 Conclusion

Although our present study did not reveal significant improvement in working memory and neurocognitive functions of patients with SCD at 1-year follow-up with hydroxyurea treatment, we identified brain regions with changes in brain activation between time points during the 2-back task using searchlight-pattern classification. More brain regions with increased BOLD signals across time points and as working memory load increased from 0- to 1-back at 1-year follow-up were found in patients with no hydroxyurea treatment. Elevated BOLD signals in patients with no hydroxyurea suggest an increase in cognitive effort during working memory processing. In contrast, patients with hydroxyurea treatment showed less changes in brain activation during working memory processing across time points and fewer brain regions with increased BOLD signals at 1-year follow-up. As working memory increased, BOLD signals in the HU group increased continuously from 0- to 2-back but BOLD signals in the non-HU group only increased from 0- to 1-back and did not increase further from 1- to 2-back. As a whole, these findings revealed the effect of hydroxyurea in maintaining work memory function from degrading over time in patients with SCD.

## Author Contributions

**JL** contributed to conceptualization, investigation, methodology, data analysis and visualization, and writing the original draft. **PZ** contributed to the study design and conduction, fMRI implementation, fMRI image analysis, data analysis, data curation, conceptualization, investigation, and methodology. **JD** contributed to investigation, conceptualization, and methodology. **TP** and **YL** contributed to statistical analysis. **MS, WW, AH, and AS** contributed to conceptualization and methodology. **KH** contributed to conceptualization and project administration. **RS** contributed to conceptualization of the study, scientific and technical direction of the analytic procedures, manuscript preparation, project administration, and supervision. All authors contributed to manuscript reading, revision, and approved the submitted version.

## Data Availability

All data produced in the present study are available upon reasonable request to the authors

## Acknowledgments

We would like to acknowledge Dr. Robert Ogg for providing their assistance in the study, and Dr. Angela J. McArthur for helping to edit the first draft.

## Data Availability Statement

The raw data supporting the conclusions of this article will be made available by the authors, without undue reservation.

## Funding

This work was supported by the Cancer Center Support (CORE) grant CA21765 from the National Cancer Institute, grant RR029005 from the National Center for Research Resources, and ALSAC. Andrew M. Heitzer was supported by K23HL166697 (National Heart, Lung, and Blood Institute) during the time of this study.

## Conflict of Interest

Dr. Akshay Sharma has received consultant fees from Spotlight Therapeutics, Medexus Inc., Vertex Pharmaceuticals, Sangamo Therapeutics, and Editas Medicine. He has also received research funding from CRISPR Therapeutics and honoraria from Vindico Medical Education. Dr. Sharma is the St. Jude Children’s Research Hospital site principal investigator of clinical trials for genome editing of sickle cell disease sponsored by Vertex Pharmaceuticals/CRISPR Therapeutics (NCT03745287), Novartis Pharmaceuticals (NCT04443907) and Beam Therapeutics (NCT05456880). The industry sponsors provide funding for the clinical trial, which includes salary support paid to Dr. Sharma’s institution. Dr. Sharma has no direct financial interest in these therapies. All other authors declare that the research was conducted in the absence of any commercial or financial relationships that could be construed as a potential conflict of interest. Dr. Andrew M. Heitzer has received consulting fees from Global Blood Therapeutics.

## Ethical approval statement

This study received ethical approval from the St. Jude Children’s Research Hospital IRB (Approval SCDMR4) on 05/26/2010.

